# Durability and determinants of anti-SARS-CoV-2 spike antibodies following the second and third doses of mRNA COVID-19 vaccine

**DOI:** 10.1101/2022.11.07.22282054

**Authors:** Shohei Yamamoto, Yusuke Oshiro, Natsumi Inamura, Takashi Nemoto, Kumi Horii, Kaori Okudera, Maki Konishi, Mitsuru Ozeki, Tetsuya Mizoue, Haruhito Sugiyama, Nobuyoshi Aoyanagi, Wataru Sugiura, Norio Ohmagari

## Abstract

**Background:** Epidemiological data regarding differences in durability and its determinants of humoral immunity following 2- and 3-dose COVID-19 vaccination are scarce.

**Methods:** We repeatedly assessed the anti-spike IgG antibody titers of 2- and 3-dose mRNA vaccine recipients among the staff of a medical and research center in Tokyo. Linear mixed models were used to estimate trajectories of antibody titers from 14 to 180 days after the last immune-conferred event (vaccination or infection) and compare antibody waning rates across prior infection and vaccination status, and across background factors in infection-naïve participants.

**Results:** A total of 6901 measurements from 2964 participants (median age, 35 years; 30% male) were analyzed. Antibody waning rate (per 30 days [95% CI]) was slower after 3-dose (25% [23–26]) than 2-dose (36% [35–37]). Participants with hybrid immunity (vaccination and infection) had further slower waning rates: 2-dose plus infection (16% [9–22]); 3-dose plus infection (21% [17–25]). Older age, male sex, obesity, coexisting diseases, immunosuppressant use, smoking, and alcohol drinking were associated with lower antibody titers, whereas these associations disappeared after 3-dose, except for sex (lower in female participants) and immunosuppressant use. Antibody waning was faster in older participants, females, and alcohol drinkers after 2-dose, whereas it did not differ after 3-dose across except sex.

**Conclusions:** The 3-dose mRNA vaccine conferred higher durable antibody titers, and previous infection further enhanced its durability. The antibody levels at a given time point and waning speed after 2-dose differed across background factors; however, these differences mostly diminished after 3-dose.

## Introduction

The widespread use of primary (1- to 2-dose) and booster (3-dose) vaccinations against coronavirus disease 2019 (COVID-19) substantially decreased the risk of severe acute respiratory syndrome coronavirus 2 (SARS-CoV-2) infection and COVID-19-related hospitalization [1, 2]. With the waning of vaccine-induced humoral immunity over time [3], however, vaccine effectiveness (VE) of 2- and 3-dose for infection has decreased [1, 2]. Understanding the duration of vaccine-induced immunity and factors accelerating the waning is critical for formulating vaccine policy, including the recommendation regarding the timing and target of an additional booster dose.

Epidemiological data are scarce regarding the waning pattern of humoral immunity following 2- and 3-dose COVID-19 vaccination and its related factors. Studies among 2-dose recipients showed that anti-SARS-CoV-2 spike antibody titers waned faster in older age [4] and female sex [3] and slower in those with a history of COVID-19 [5], but no such investigation has been done for 3-dose. It remains elusive for both doses whether the waning speed of antibody differs depending on obesity [6, 7], comorbid conditions [7, 8], immunosuppression [7, 8], or behavioral factors (smoking and alcohol drinking [9-11]), which have been linked to lower immune response to COVID-19 vaccine.

Here we assess the long-term humoral response and its determinants following 2- and 3-dose COVID-19 mRNA vaccines and compare the waning rates between 2- and 3-dose recipients using the data of a repeat serological study among the staff of a national center for medical care and research in Tokyo.

## Methods

### Study setting

National Center for Global Health and Medicine, Japan (NCGM) is a medical research center for specific areas, including infectious diseases. As a mission, NCGM has played a major role in the care and research of COVID-19 since the early stage of the epidemic [12] and has accepted many inpatients with severe COVID-19. The in-house vaccination program using COVID-19 mRNA-LNP BNT162b2 (Pfizer-BioNTech) was conducted from March to June 2021 for the first and second doses and from December 2021 to February 2022 for the third vaccination.

In the NCGM, a repeat serological study was launched in July 2020 to monitor the spread of SARS-CoV-2 infection among staff during the COVID-19 pandemic [13]. As of June 2022, we have completed six surveys: the first (July 2020), second (October to December 2020), third (June 2021), fourth (December 2021), fifth (March 2022), and sixth (June 2022). In each survey, we measured anti-SARS-CoV-2 nucleocapsid- and spike- (from the second survey onward) protein antibodies using Abbott and Roche assays and collected patient information, including histories of SARS-CoV-2 vaccination and infection, body composition, morbid status, and behavioral factors. Self-reported vaccination status was validated using the information provided by the NCGM Labor Office, whereas the self-reported history of SARS-CoV-2 infection was validated against COVID-19 patient records maintained by the NCGM Hospital Infection Prevention and Control Unit. Written informed consent was obtained from all participants. This study was approved by the NCGM ethics committee (approval number: NCGM-G-003598).

### Analytic samples

We retrieved 7,340 antibody measurements from 3,151 staff who participated in at least one of the third to sixth surveys more than seven days after receiving two or three doses of mRNA COVID-19 vaccines (**Supplemental Figure 1**). Of these, we excluded 301 measurements that tested seropositive with anti-SARS-CoV-2 nucleocapsid (N) protein assays (Abbott and/or Roche) without a history of SARS-COV-2 infection because the timing of infection could not be determined (**Supplemental Figure 2**). Of the remaining data, we further excluded 138 measurements missing any of the following variables: body mass index (n=80), coexisting diseases (n=74), immunosuppressive drug use (n=41), smoking status (n=103), and alcohol drinking status (n=48), leaving 6,901 measurements of 2,964 participants for the analysis of antibody waning by vaccine and infection status. Of these, we excluded 325 data tested seropositive with anti-N assays and/or had a history of SARS-CoV-2 infection, leaving 6,576 measurements from 2,906 participants for the association analyses among infection-naïve participants.

### Antibody testing

At each survey, we quantitatively measured the levels of antibodies against the receptor-binding domain of the SARS-CoV-2 spike protein using the AdviseDx SARS-CoV-2 IgG II assay (Abbott) (immunoglobulin [Ig] G [IgG]). We also qualitatively measured antibodies against the SARS-CoV-2 N protein using the SARS-CoV-2 IgG assay (Abbott) and Elecsys^®^ Anti-SARS-CoV-2 RUO (Roche).

### Stratifying variables

We obtained information on age and sex from the NCGM Labor Office and height, weight, coexisting diseases, immunosuppressant use, tobacco products use, and alcohol use via a questionnaire at the survey when each participant was first included as analytic samples. We calculated body mass index (BMI) as the weight (kg) divided by the square of height (m) and defined obesity as ≥27.5 kg/m^2^ of BMI according to the WHO classification for Asians [14]. Coexisting diseases were defined if a participant reported having at least one of the following diseases: hypertension, diabetes, chronic kidney disease, dyslipidemia, cancer, and cardiovascular disease. Immunosuppressant use was defined if a participant reported using steroids (except topical or inhaled), immunosuppressants, or anticancer drugs. Current smokers were defined as users of tobacco products, including conventional cigarettes or heated tobacco products. Regular alcohol use was defined as those who consumed alcoholic beverages at least once per week.

### Statistical analysis

Trajectories of the antibody titers over time were estimated by a multivariable mixed-effects model with a person-specific random intercept and slope. The dependent variable was log-transformed antibody titers. Fixed effect covariates included time since last exposure to SARS-CoV-2 infection or vaccination (continuous and squared terms), vaccination status (2- or 3-dose), history of COVID-19 (yes or no), sex (male or female), age (<40 or ≥40 years), BMI (<27.5 or ≥27.5 kg/m^2^), coexisting diseases (no or yes), immunosuppressant use (no or yes), use of tobacco products (no or current), and regular alcohol use (no or yes). Interaction terms between time, vaccination status, and each of those covariates were also included as fixed effects to estimate the decline slope of antibodies following 2- and 3-dose for each subgroup and to compare the slope across each subgroup in 2- and 3-dose recipients. The estimated effects of covariates were back-transformed and presented as ratios of geometric means and geometric mean titers (GMTs) with 95% confidence intervals (CIs). Using the post-estimation command “*margins*” in Stata, we estimated the GMT of spike antibodies at Day 14, 30, 90, and 180 for each subgroup. Results for groups that detected statistical significance in the comparison of antibody slopes were visualized using the Stata command “*marginsplot*.” We repeated the analysis to examine the differences in waning rates across patterns of immune-conferring events (i.e., vaccination before or after SARS-CoV-2 infection) and across timing of infection (pre-Omicron or Omicron BA.1/BA.2 predominant waves). All statistical analyses were performed using Stata 17.0 (StataCorp, College Station, TX). All P values were 2-sided, and P < 0.05 was considered statistically significant.

## Results

### Characteristics of participants

The median age of study participants was 35 (interquartile range [IQR]: 27–47), 30.2% were male, and major occupation was nurse (37.2%), doctor (18.3%), allied health professional (13.7%), or administrative staff (9.6%) (**Table 1**). Those with obesity, coexisting diseases, and immunosuppression were 5.5%, 11.1%, and 1.8%, respectively, and tobacco product users and regular drinkers were 7.7% and 37.6%, respectively. The number of participants who attended surveys one to four times was 870 (29.4%), 755 (25.5%), 835 (28.2%), and 504 (17.0%), respectively.

**Table 1.**
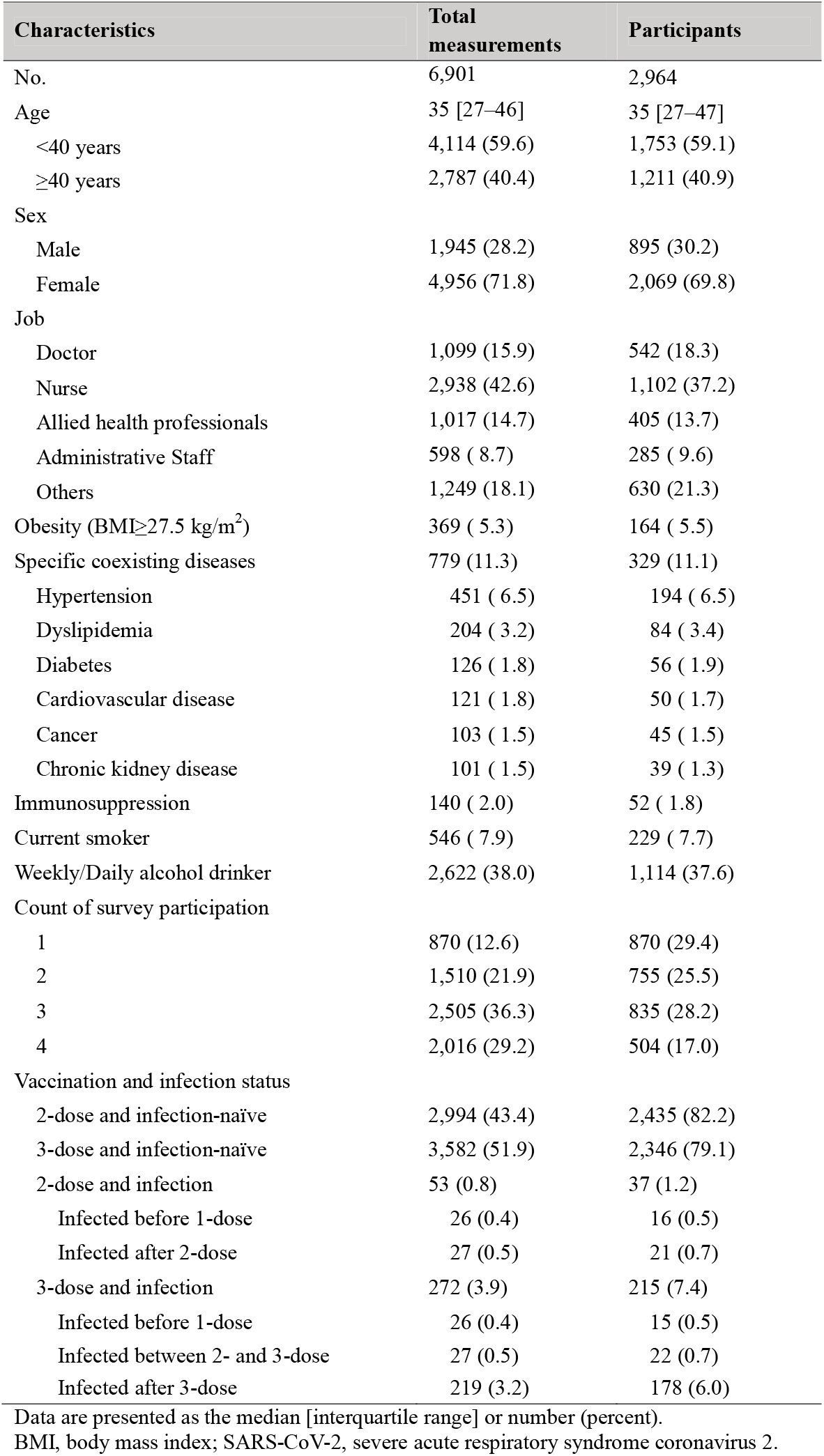
Characteristics of total measurements and participants

Of 6,901 total measurements, data of infection-naïve participants following 2- and 3-dose were 2,994 (43.4%) and 3,582 (51.9%), respectively. Those who received 2-dose with a history of SARS-CoV-2 infection were 53 (0.8%), including those who were infected before 1-dose (n=26) and infected after 2-dose (n=27). The 3-dose recipients with infection history were 272, including those who were infected before 1-dose (n=26), infected between 2- and 3-dose (n=27), and infected after 3-dose (n=219). The median intervals (min–max) from the last immune-conferring event (infection or vaccination) to the blood sampling among 2- and 3-dose recipients were 68 days (8–462) and 122 days (8–221), respectively (**Supplemental Figure 3**).

### Waning of antibody titers across vaccination and infection status

Among the infection-naïve participants, the waning rate of antibody titers was 17% (95% CIs: 16–19) slower after the 3-dose (25% per 30 days) compared with 2-dose (36% per 30 days) (**Figure 1** and **Table 2**). The difference in antibody titers between 3-dose and 2-dose became larger over time: estimated ratios of GMT (95% CIs) of 3-dose to 2-dose on days 14, 30, 90, and 180 were 1.70 (1.59–1.83), 1.98 (1.88–2.08), 3.13 (3.02–3.25), and 4.52 (4.29–4.77), respectively.

**Figure 1.**
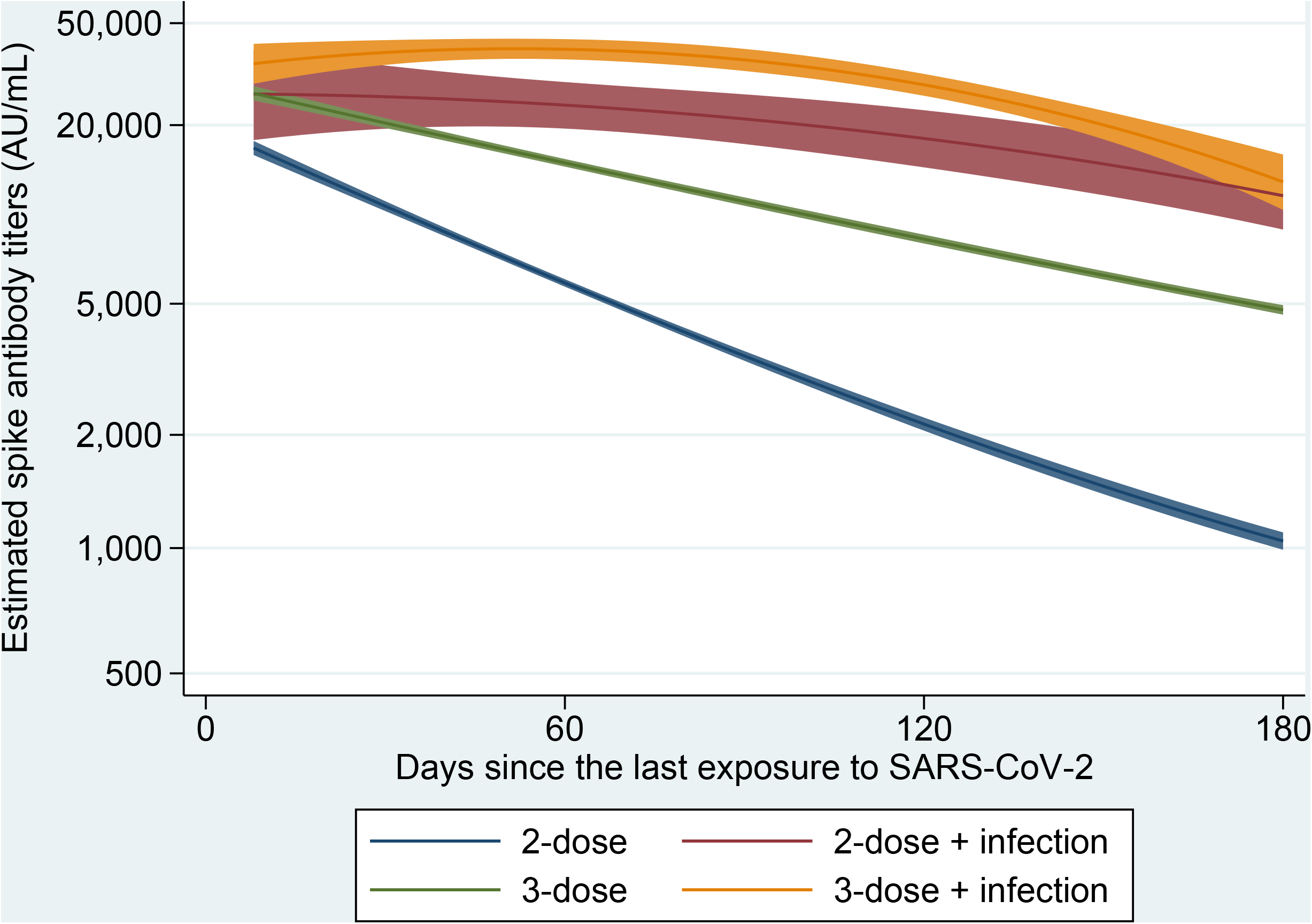
Waning of anti-SARS-CoV-2 spike protein antibody titers from the last exposure to SARS-CoV-2 (infection or vaccination) Curves are shown the estimated trajectories adjusted for age (<40 or ≥40 years), sex (male or female), body mass index (<27.5 or ≥27.5 kg/m^2^), coexisting diseases (yes or no), immunosuppressant use (yes or no), smoking status (current or non-smoker), and frequency of alcohol drinking (<1 or ≥1 time/week). A blue line shows participants who received 2-dose with infection-naïve (N=2994), while a red line shows those who received 2-dose with a history of SARS-CoV-2 infection (N=53). A green line shows those who received 3-dose with infection-naïve (N=3582), and an orange line shows those who received 3-dose with a history of the infection (N=272). Solid lines are the estimated geometric mean of anti-SARS-CoV-2 spike protein antibody titers, and shaded areas are the corresponding 95% confidence intervals. N, number of measurements; SARS-CoV-2, severe acute respiratory syndrome coronavirus 2.

**Table 2.** Waning of antibody titers since the last immune-conferring event (vaccination or infection) among 2- or 3-dose vaccine recipients

|  | Estimated spike antibody titers (AU/mL) following the last immune-conferring event, GMT (95% CI) |  |  |  | Slope of antibody waning, RoM (95%CI) |  |  |
| --- | --- | --- | --- | --- | --- | --- | --- |
|  | Day 14 | Day 30 | Day 90 | Day 180 | Per 30 days | Comparison of slopes |  |
| 2-dose | 14426 (13692–15199) | 10284 (9893–10690) | 3393 (3282–3509) | 1034 (983–1089) | 0.64<br>(0.63–0.65) | reference | – |
| 3-dose | 24389 (23069–25783)* | 20280 (19450–21146)* | 10663 (10317–11021)* | 4699 (4540–4864)* | 0.75<br>(0.74–0.76) | 1.17<br>(1.16–1.19) | reference |
| 2-dose +<br>infection | 25904 (18164–36942)* | 25400 (19323–33388)* | 20995 (17000–25928)*† | 11167 (8565–14560)*† | 0.84<br>(0.78–0.91) | 1.32<br>(1.23–1.43) | 1.13<br>(1.05–1.21) |
| 3-dose +<br>infection | 35131 (30078–41032)*† | 37543 (33590–41961)*† | 35119 (31821–38758)*† | 12476 (9986–15586)*† | 0.79<br>(0.75–0.83) | 1.24<br>(1.17–1.31) | 1.05<br>(1.01–1.11) |

Shown are the estimated GMT of anti-SARS-CoV-2 spike protein antibody titers on 14, 30, 90, and 180 days since the last immune-conferring event (vaccination or infection) and the estimated RoM of antibody waning slopes, with adjustment for age (<40 or ≥40 years), sex (male or female), body mass index (<27.5 or ≥27.5 kg/m<sup>2</sup>), coexisting diseases (yes or no), immunosuppressant use (yes or no), smoking status (current or non-smoker), and frequency of alcohol drinking (<1 or ≥1 time/week).
\*: P<0.05 (reference is 2-dose)
†: P<0.05 (reference is 3-dose)
GMT, geometric mean titer; RoM, ratio of means

The 2- and 3-dose recipients with a history of SARS-CoV-2 infection had slower antibody waning rates than their infection-naïve counterparts (**Figure 1** and **Table 2**). Among the 2-dose recipients, those with a history of infection had consistently higher antibody titers during days 14 to 180 and a 32% (95%CIs: 23–43) slower waning rate than those without infection. Among the 3-dose recipients, those with a history of infection had higher antibody titers during days 14 to 180 and a 5% (95%CIs: 1–11) slower decline rate than those without infection. The 2-dose recipients with infection had higher antibody titers and a 13% (95%CIs: 5–21) slower waning rate than 3-dose recipients without infection. Among those with a history of infection, there were no material differences in antibody waning rates across sequence patterns of immune-conferring events (vaccination before or after infection) and timing of infection (i.e., pre-Omicron or Omicron BA.1/BA.2 predominant waves) (**Supplemental Table 1 and 2**).

### Waning of antibody titers across background factors

**Table 3** and **Table 4** show the patterns of antibody waning following 2- and 3-dose vaccines, respectively, according to background factors among infection-naïve participants. Those aged 40 years or older had consistently lower antibody titers during days 14 to 180 and had a 4% (95%CIs: 1–7) faster antibody waning rate than those aged less than 40 years among 2-dose recipients, whereas such age difference in antibody waning was not observed among 3-dose recipients (**Figure 2A**). Females had higher antibody titers than males after 2-dose recipients, whereas the opposite was observed after 3-dose. Antibody declined 3% faster in females than males among both 2-dose (ratio of slopes: 0.97, 95%CIs: 0.94–0.99) and 3-dose recipients (ratio of slopes: 0.97, 95%CIs: 0.95–0.99) (**Figure 2B**). Obesity was associated with lower antibody titers in 2-dose recipients but not in 3-dose recipients. Waning rates were similar in those with and without obesity after 2- or 3-dose. Coexisting diseases were associated with lower antibody titers among 2-dose recipients but not among 3-dose recipients. Antibody waning rates among 2- and 3-dose recipients were similar irrespective of coexisting disease status. Those using immunosuppressants had lower antibody titers following 2- and 3-dose than those without, while there were no differences in the waning rates by immunosuppressant use. Current users of tobacco products had significantly lower antibody titers after 2-dose than non-smokers, but the difference became smaller and statistically not significant after 3-dose. Antibody waning rates were similar irrespective of smoking status among 2- and 3-dose recipients. Regular alcohol drinkers had lower antibody titers following 2-dose than non-drinkers or occasional drinkers, whereas such difference was not observed after 3-dose. Waning rates were 3% faster in regular alcohol drinkers than non-drinker/occasional-drinkers among 2-dose recipients (ratio of slopes: 0.97, 95%CIs: 0.94–0.99), while their slopes were similar among 3-dose recipients (**Figure 3C**).

**Table 3.** Characteristics of antibody waning following the second vaccination across the background factors among infection-naïve participants.

|  |  | Estimated spike antibody titers (AU/mL) following 2-dose, GMT (95% CI) |  |  |  | Slope of antibody waning, RoM (95%CI) |  |
| --- | --- | --- | --- | --- | --- | --- | --- |
| Characteristics |  | Day 14 | Day 30 | Day 90 | Day 180 | Per 30 days | Comparison of slopes |
| Age | <40 years | 16076<br>(15103–17113) | 11618<br>(11074–12189) | 3967<br>(3803–4137) | 1210<br>(1140–1285) | 0.64<br>(0.63–0.65) | reference |
|  | ≥40 years | 13405<br>(12187–14744)* | 9137<br>(8538–9778)* | 2651<br>(2502–2809)* | 749<br>(677–829)* | 0.61<br>(0.60–0.63) | **0.96**<br>**(0.93–0.99)** |
| Sex | Male | 11947<br>(10886–13112) | 8685<br>(8098–9314) | 3041<br>(2865–3228) | 973<br>(885–1070) | 0.65<br>(0.64–0.67) | reference |
|  | Female | 15632<br>(14706–16617)* | 11078<br>(10588–11591)* | 3585<br>(3446–3729)* | 1070<br>(1008–1135) | 0.63<br>(0.62–0.64) | **0.97**<br>**(0.94–0.99)** |
| Obesity | <27.5 kg/m<sup>2</sup> | 14508<br>(13765–15291) | 10376<br>(9977–10791) | 3450<br>(3335–3569) | 1050<br>(997–1106) | 0.64<br>(0.63–0.65) | reference |
|  | ≥27.5 kg/m<sup>2</sup> | 15147<br>(11122–20627) | 9956<br>(8081–12266) | 2690<br>(2257–3206)* | 827<br>(622–1101) | 0.62<br>(0.57–0.67) | 0.97<br>(0.89–1.05) |
| Coexisting diseases | No | 14786<br>(14077–11086) | 10583<br>(10167–11017) | 3525<br>(3404–3650) | 1071<br>(1016–1129) | 0.64<br>(0.63–0.65) | reference |
|  | Yes | 14077<br>(11086–17875) | 9350<br>(7977–10959) | 2572<br>(2259–2928)* | 764<br>(621–941)* | 0.62<br>(0.58–0.65) | 0.96<br>(0.91–1.02) |
| Immunosuppressant use | No | 14564<br>(13825–15343) | 10388<br>(9993–10798) | 3432<br>(3319–3549) | 1046<br>(993–1101) | 0.64<br>(0.63–0.65) | reference |
|  | Yes | 8074<br>(5221–12486)* | 6315<br>(4588–8694)* | 2681<br>(2108–3409)* | 897<br>(595–1352) | 0.68<br>(0.61–0.76) | 1.06<br>(0.95–1.20) |
| Smoking | Non-smokers | 14663<br>(13881–15490) | 10463<br>(10048–10894) | 3459<br>(3340–3582) | 1052<br>(998–1110) | 0.64<br>(0.63–0.65) | reference |
|  | Current smokers | 11911<br>(10084–14069)* | 8606<br>(7563–9793)* | 2957<br>(2642–3309)* | 930<br>(780–1108) | 0.65<br>(0.62–0.68) | 1.01<br>(0.97–1.06) |
| Alcohol drinking | Non or Occasional | 14818<br>(13926–15768) | 10712<br>(10220–11228) | 3671<br>(3522–3826) | 1134<br>(1065–1207) | 0.64<br>(0.63–0.65) | reference |
|  | Weekly or Daily | 14269<br>(12962–15706) | 9868<br>(9212–10572) | 2998<br>(2830–3176)* | 884<br>(807–968)* | 0.62<br>(0.61–0.64) | **0.97**<br>**(0.94–0.99)** |

Shown are the estimated GMT of anti-SARS-CoV-2 spike protein antibody titers on 14, 30, 90, and 180 days since the second mRNA vaccination and the estimated RoM of antibody waning slopes, with adjustment for age ( $<40$ or $\geq 40$ years), sex (male or female), body mass index ( $<27.5$ or $\geq 27.5$ kg/m<sup>2</sup>), coexisting diseases (yes or no), immunosuppressant use (yes or no), smoking status (current or non-smoker), and frequency of alcohol drinking ( $<1$ or $\geq 1$ time/week).
\*P<0.05
GMT, geometric mean titer; RoM, ratio of means

**Table 4.** Characteristics of antibody waning following the third vaccination across the background factors among infection-naïve participants.

|  |  | Estimated spike antibody titers (AU/mL) following 3-dose, GMT (95% CI) |  |  |  | Slope of antibody waning, RoM (95%CI) | Comparison of slopes |
| --- | --- | --- | --- | --- | --- | --- | --- |
| Characteristics |  | Day 14 | Day 30 | Day 90 | Day 180 | Per 30 days |  |
| Age | <40 years | 25788<br>(24167–27517) | 21110<br>(20077–22196) | 10696<br>(10242–11171) | 4756<br>(4543–4980) | 0.75<br>(0.74–0.75) | reference |
|  | ≥40 years | 23286<br>(20993–25829) | 19566<br>(18132–21114) | 10517<br>(9992–11069) | 4552<br>(4323–4794) | 0.75<br>(0.74–0.76) | 1.00<br>(0.99–1.02) |
| Sex | Male | 25563<br>(22422–29144) | 22274<br>(20255–24494) | 13168<br>(12360–14029) | 5827<br>(5472–6204) | 0.76<br>(0.75–0.78) | reference |
|  | Female | 24200<br>(22803–25683) | 19770<br>(18889–20692)* | 9926<br>(9558–10309)* | 4336<br>(4166–4513)* | 0.74<br>(0.73–0.75) | **0.97**<br>**(0.95–0.99)** |
| Obesity | <27.5 kg/m<sup>2</sup> | 24455<br>(23117–25870) | 20284<br>(19442–21161) | 10601<br>(10251–10963) | 4678<br>(4517–4845) | 0.75<br>(0.74–0.76) | reference |
|  | ≥27.5 kg/m<sup>2</sup> | 27046<br>(20590–35526) | 23099<br>(18859–28291) | 12810<br>(11073–14820)* | 5323<br>(4630–6120) | 0.75<br>(0.72–0.78) | 1.00<br>(0.96–1.04) |
| Coexisting diseases | No | 24632<br>(23250–26096) | 20342<br>(19472–21250) | 10528<br>(10168–10901) | 4668<br>(4503–4839) | 0.75<br>(0.74–0.75) | reference |
|  | Yes | 24091<br>(20053–28942) | 20814<br>(18166–23850) | 11966<br>(10833–13216) | 5133<br>(4619–5704) | 0.76<br>(0.74–0.78) | 1.01<br>(0.98–1.04) |
| Immunosuppressant use | No | 24737<br>(23391–26160) | 20554<br>(19708–21437) | 10788<br>(10436–11151) | 4757<br>(4596–4923) | 0.75<br>(0.74–0.76) | reference |
|  | Yes | 17168<br>(12090–24379)* | 14067<br>(10717–18465)* | 7065<br>(5580–8945)* | 2987<br>(2331–3828)* | 0.74<br>(0.69–0.78) | 0.98<br>(0.93–1.04) |
| Smoking | Non-smokers | 24621<br>(23232–26092) | 20479<br>(19606–21392) | 10777<br>(10416–11150) | 4750<br>(4584–4922) | 0.75<br>(0.74–0.76) | reference |
|  | Current smokers | 23542<br>(19672–28175) | 19415<br>(16893–22313) | 9959<br>(8824–11239) | 4309<br>(3829–4848) | 0.74<br>(0.72–0.77) | 0.99<br>(0.96–1.02) |
| Alcohol drinking | Non or Occasional | 25174<br>(23477–26994) | 20807<br>(19741–21931) | 10814<br>(10372–11275) | 4839<br>(4632–5056) | 0.75<br>(0.74–0.76) | reference |
|  | Weekly or Daily | 23513<br>(21483–25735) | 19708<br>(18411–21097) | 10509<br>(9961–11087) | 4513<br>(4272–4769) | 0.75<br>(0.74–0.76) | 1.00<br>(0.98–1.01) |

Shown are the estimated GMT of anti-SARS-CoV-2 spike protein antibody titers on 14, 30, 90, and 180 days since the third mRNA vaccination and the estimated RoM of antibody waning slopes, with adjustment for age ( $<40$ or $\geq 40$ years), sex (male or female), body mass index ( $<27.5$ or $\geq 27.5$ kg/m<sup>2</sup>), coexisting diseases (yes or no), immunosuppressant use (yes or no), smoking status (current or non-smoker), and frequency of alcohol drinking ( $<1$ or $\geq 1$ time/week).

**Figure 2.**
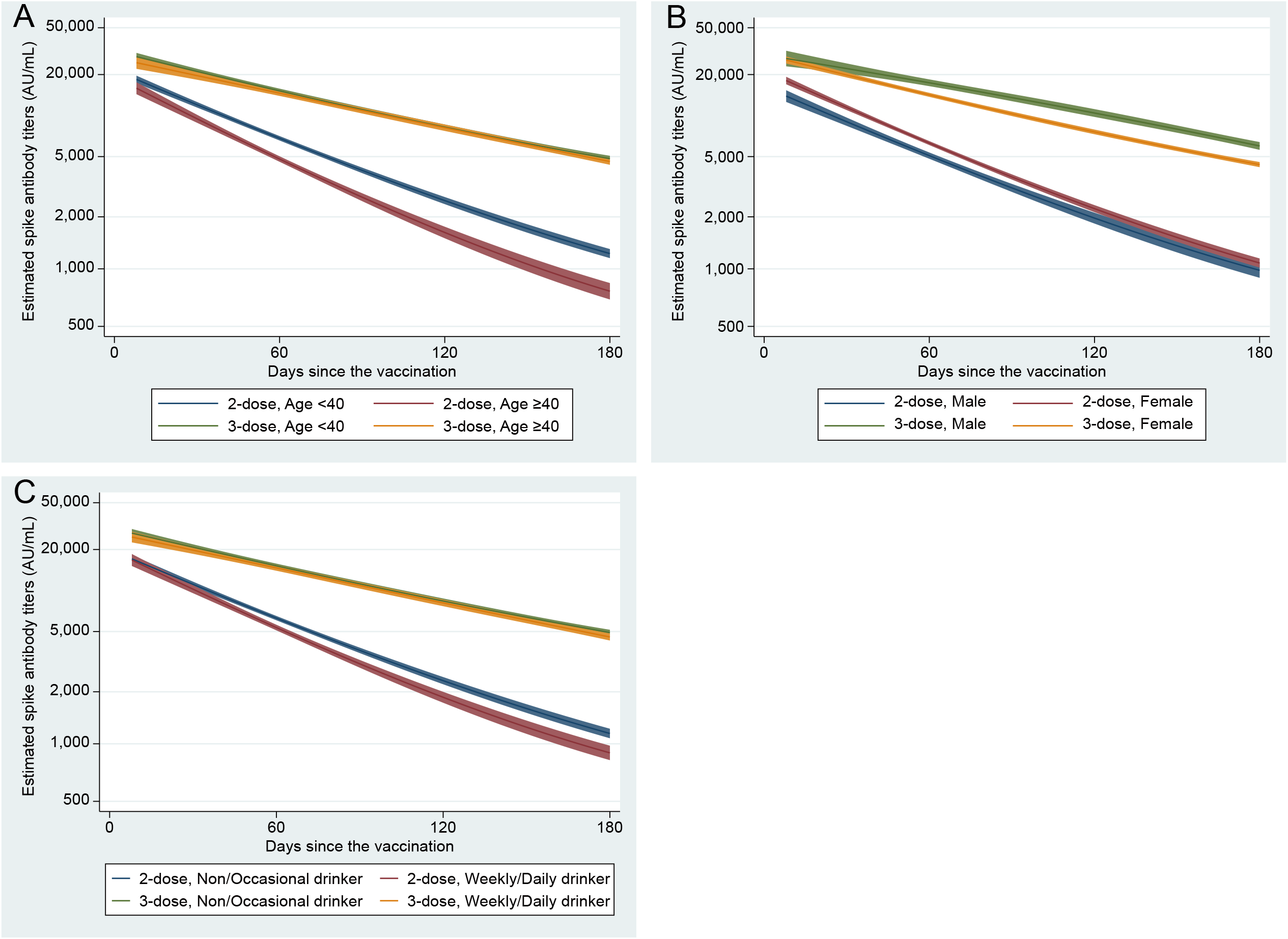
Trajectories of anti-SARS-CoV-2 spike protein antibody titers following 2- and 3-dose vaccination by age (**A**), sex (**B**), and alcohol drinking status (**C**) among infection-naïve participants Curves are shown the estimated trajectories for individuals who had received 2-dose (blue) and those who had received 3-dose (red) adjusted for age (<40 or ≥40 years), sex (male or female), body mass index (<27.5 or ≥27.5 kg/m^2^), coexisting diseases (yes or no), immunosuppressant use (yes or no), smoking status (current or non-smoker), and frequency of alcohol drinking (<1 or ≥1 time/week). Solid lines are the estimated geometric mean of anti-SARS-CoV-2 spike protein antibody titers, and shaded areas are the corresponding 95% confidence intervals.

## Discussion

In the analysis of repeat serosurvey data among recipients of COVID-19 mRNA vaccines, we found that the durability of antibody titers following 3-dose was higher than following 2-dose and a history of SARS-CoV-2 infection further enhanced the sustainability of antibody titers over time. Older age, male sex, obesity, coexisting diseases, immunosuppressant use, use of tobacco products, and alcohol drinking were each associated with lower antibody titers after 2-doses, whereas these associations disappeared after 3-dose, except for sex (females became lower) and immunosuppressant use (still lower). Antibody waning rates were faster in older staff, females, and alcohol drinkers after 2-dose, and still faster in females after 3-dose. Obesity, coexisting diseases, immunosuppressant use, and tobacco product use were not associated with the waning speed of antibodies after 2- and 3-dose.

Our findings that the higher durability of antibody titers after the 3-dose of mRNA vaccine than after 2-dose is consistent with a previous study of Israeli healthcare workers [3] and compatible with the effectiveness studies of mRNA vaccines showing a slower waning of VE for infection over time in recipients of 3-dose relative to those of 2-dose [15, 16]. In addition, we found that hybrid immunity (vaccination and infection) has added durability over the immunity induced by vaccination alone. The novelty of this study is that 3-dose plus infection has led to more durable antibodies than 3-dose alone, which is compatible with a study reporting a higher VE against infection for 3-dose plus prior infection versus 3-dose alone [16]. Interestingly, we observed that those with 2-dose plus infection had more durable antibody titers than those with 3-dose alone. This result suggests that natural infection conferred more durable immunity than vaccine alone and is compatible with the results of a meta-analysis of VE studies [16, 17].

Among the individuals with hybrid immunity, there were no material differences in the durability of antibody titers across sequence patterns of hybrid immunity (vaccination before or after infection) and timing of the infected phase (pre-Omicron or Omicron BA.1/BA.2 dominant waves). Previous studies have shown that neither patterns of hybrid immunity [18] nor types of infected variants [19] affect the cross-sectional levels of humoral immunity against the ancestral strain. Our studies expanded the literature by showing that such differences do not affect the longevity of humoral immunity as well.

Among the infection-naïve participants, we observed that older age, male sex, obesity, coexisting diseases, immunosuppressant use, smoking, and regular alcohol use were each associated with lower antibody titers throughout 14 to 180 days following the second dose. These results were consistent with previous cross-sectional studies [6-10]. After the 3-dose, however, these associations disappeared except for sex and immunosuppressant use, suggesting that a booster shot can enhance immunity and minimize the difference in immunity levels between groups with different backgrounds. Still, those with immunosuppressive status had lower antibody titers after 3-dose, a finding compatible with the VE study among 2- and 3-dose vaccine recipients [2]. Our results highlight the need for careful monitoring of infection risk and consideration of additional boosters among individuals receiving immunosuppressants.

Regarding antibody waning over time, we observed a faster decline associated with older age, female sex, and regular alcohol use after 2-dose (3–4%). After 3-dose, the differences by age and alcohol used disappeared, whereas the female sex remained a significant driver of antibody decline (3%). This result suggests that a booster dose plays a role in eliminating gaps in antibody durability by background factors. We have no plausible explanation for the faster waning in females after both 2- and 3-doses. Interestingly, a meta-analysis of clinical trials on primary (two doses) COVID-19 vaccines showed that females have a significantly lower VE than males [20]; nonetheless, subsequence studies on VE after 3-dose and waning of VE after 2- and 3-dose had not examined any sex difference. Our results highlight the need for further studies regarding sex-difference in vaccine efficacy and immunogenicity.

This study has some strengths. First, this well-designed longitudinal data set with detailed information on demographic, clinical, and behavioral factors and repeated anti-spike antibody measurements allowed us to investigate the trajectories of vaccine and infection-induced antibody titers over time across these factors. Second, we also repeatedly assessed anti-N antibodies; thus, we were able to exclude the undiagnosed suspected COVID-19 patients from the infection-naïve group. Several limitations should be acknowledged. First, we did not measure neutralizing antibodies, a more reliable marker of the humoral immune response. Nevertheless, spike IgG antibody titers measured with the assay we employed (Abbott) are reported to correlate well with live-virus neutralizing antibody titers against Ancestral, Delta, and Omicron variants (Spearman’s ρ=0.91, 0.83, and 0.49, respectively) [21]. Second, we did not assess cellular immune response, another key mechanism of infection protection [22]. Third, the present participants were mainly healthcare workers and relatively younger and healthier than the general population, potentially limiting the generalizability of our results.

## Conclusion

Among the recipients of 2- and 3-dose mRNA COVID-19 vaccines, the waning of anti-SARS-CoV-2 spike antibody titers over time was more durable after 3-dose than 2-dose, and hybrid immunologic status (vaccination and infection) conferred additional longevity of antibody titers. Older age, female sex, and alcohol drinking were each associated with a faster decline of antibody titers after 2-dose, whereas only female sex remained a significant factor after 3-dose. These results inform policymakers of the importance of consideration for personal background, the number of vaccinations received, and the history of COVID-19 infection in formulating vaccine recommendations.

## Data Availability

All data produced in the present study are available upon reasonable request to the authors.

## Acknowledgements

We thank Mika Shichishima for her contribution to data collection and the staff of the Laboratory Testing Department for their contribution to measuring antibody testing.

## References

1. Feikin DR, Higdon MM, Abu-Raddad LJ, et al. Duration of effectiveness of vaccines against SARS-CoV-2 infection and COVID-19 disease: results of a systematic review and meta-regression. The Lancet 2022; 399(10328): 924–44.

2. Tseng HF, Ackerson BK, Luo Y, et al. Effectiveness of mRNA-1273 against SARS-CoV-2 Omicron and Delta variants. Nat Med 2022; 28(5): 1063–71.

3. Gilboa M, Regev-Yochay G, Mandelboim M, et al. Durability of Immune Response After COVID-19 Booster Vaccination and Association With COVID-19 Omicron Infection. JAMA Network Open 2022; 5(9): e2231778.

4. Kim HJ, Yun HJ, Kim J, Kym S, Choi Q. Antibody response to second dose of the BNT162b2 mRNA vaccine in the first 12 weeks in South Korea: A prospective longitudinal study. Vaccine 2022; 40(3): 437–43.

5. Zhong D, Xiao S, Debes AK, et al. Durability of Antibody Levels After Vaccination With mRNA SARS-CoV-2 Vaccine in Individuals With or Without Prior Infection. JAMA 2021; 326(24): 2524.

6. Yamamoto S, Mizoue T, Tanaka A, et al. Sex-associated differences between BMI and SARS-CoV-2 antibody titers following the BNT162b2 vaccine. Obesity 2022; 30(5): 999–1003.

7. Yamamoto S, Tanaka A, Oshiro Y, Inamura N, Mizoue T, Ohmagari N. Antibody responses and correlates after two and three doses of BNT162b2 COVID-19 vaccine. Infection 2022.

8. Levin EG, Lustig Y, Cohen C, et al. Waning Immune Humoral Response to BNT162b2 Covid-19 Vaccine over 6 Months. N Engl J Med 2021.

9. Kageyama T, Ikeda K, Tanaka S, et al. Antibody responses to BNT162b2 mRNA COVID-19 vaccine and their predictors among healthcare workers in a tertiary referral hospital in Japan. Clin Microbiol Infect 2021.

10. Nomura Y, Sawahata M, Nakamura Y, et al. Age and Smoking Predict Antibody Titres at 3 Months after the Second Dose of the BNT162b2 COVID-19 Vaccine. Vaccines 2021; 9(9): 1042.

11. Yamamoto S, Tanaka A, Ohmagari N, et al. Use of heated tobacco products, moderate alcohol drinking, and anti-SARS-CoV-2 IgG antibody titers after BNT162b2 vaccination among Japanese healthcare workers. Prev Med 2022; 161: 107123.

12. Hayakawa K, Kutsuna S, Kawamata T, et al. SARS-CoV-2 infection among returnees on charter flights to Japan from Hubei, China: a report from National Center for Global Health and Medicine. Glob Health Med 2020; 2(2): 107–11.

13. Yamamoto S, Maeda K, Matsuda K, et al. Coronavirus Disease 2019 (COVID-19) Breakthrough Infection and Post-Vaccination Neutralizing Antibodies Among Healthcare Workers in a Referral Hospital in Tokyo: A Case-Control Matching Study. Clin Infect Dis 2021.

14. Appropriate body-mass index for Asian populations and its implications for policy and intervention strategies. The Lancet 2004; 363(9403): 157–63.

15. Andrews N, Stowe J, Kirsebom F, et al. Covid-19 Vaccine Effectiveness against the Omicron (B.1.1.529) Variant. N Engl J Med 2022; 386(16): 1532–46.

16. Altarawneh HN, Chemaitelly H, Ayoub HH, et al. Effects of Previous Infection and Vaccination on Symptomatic Omicron Infections. N Engl J Med 2022; 387(1): 21–34.

17. Bobrovitz N, Ware H, Ma X, et al. Protective effectiveness of prior SARS-CoV-2 infection and hybrid immunity against Omicron infection and severe disease: a systematic review and meta-regression. medRxiv (preprint) 2022.

18. Bates TA, McBride SK, Leier HC, et al. Vaccination before or after SARS-CoV-2 infection leads to robust humoral response and antibodies that effectively neutralize variants. Science Immunology 2022; 7(68): eabn8014.

19. Suryawanshi R, Ott M. SARS-CoV-2 hybrid immunity: silver bullet or silver lining? Nature Reviews Immunology 2022; 22(10): 591–2.

20. Bignucolo A, Scarabel L, Mezzalira S, Polesel J, Cecchin E, Toffoli G. Sex Disparities in Efficacy in COVID-19 Vaccines: A Systematic Review and Meta-Analysis. Vaccines 2021; 9(8): 825.

21. Servellita V, Syed AM, Morris MK, et al. Neutralizing immunity in vaccine breakthrough infections from the SARS-CoV-2 Omicron and Delta variants. Cell 2022; 185(9): 1539-48.e5.

22. Tan AT, Linster M, Tan CW, et al. Early induction of functional SARS-CoV-2-specific T cells associates with rapid viral clearance and mild disease in COVID-19 patients. Cell Rep 2021; 34(6): 108728.

