## Supplementary materials for "Durability and determinants of anti-SARS-CoV-2 spike antibodies following the second and third doses of mRNA COVID-19 vaccine"

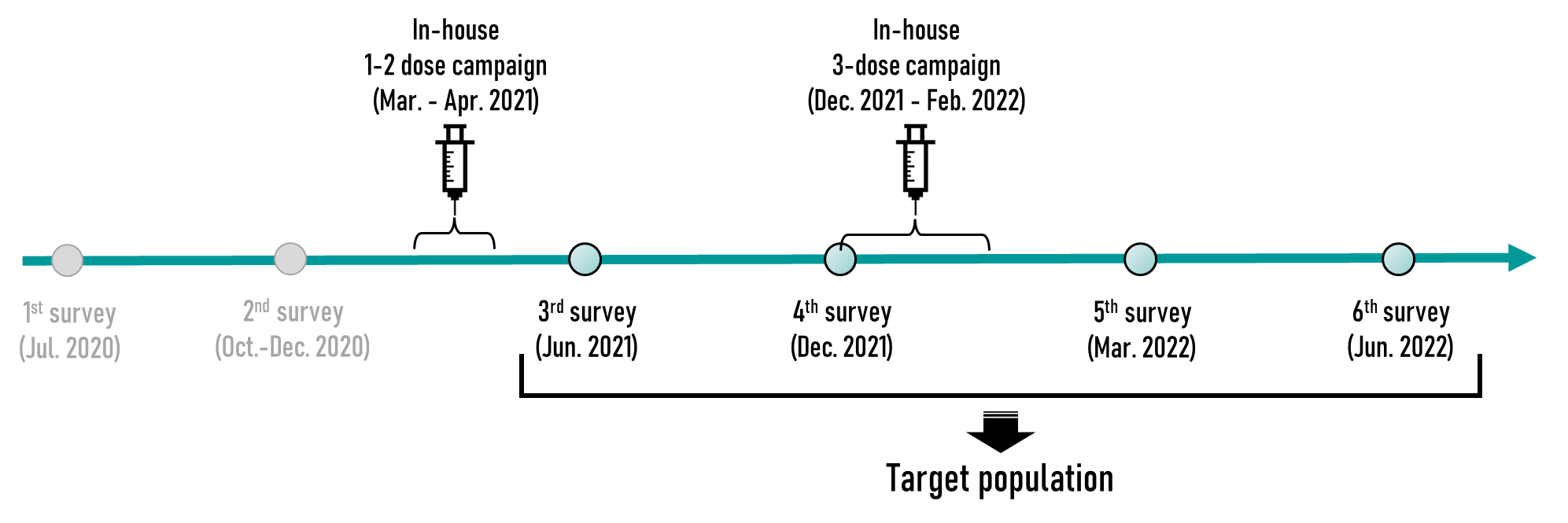

**Supplemental Figure 1.** Study design and participants.

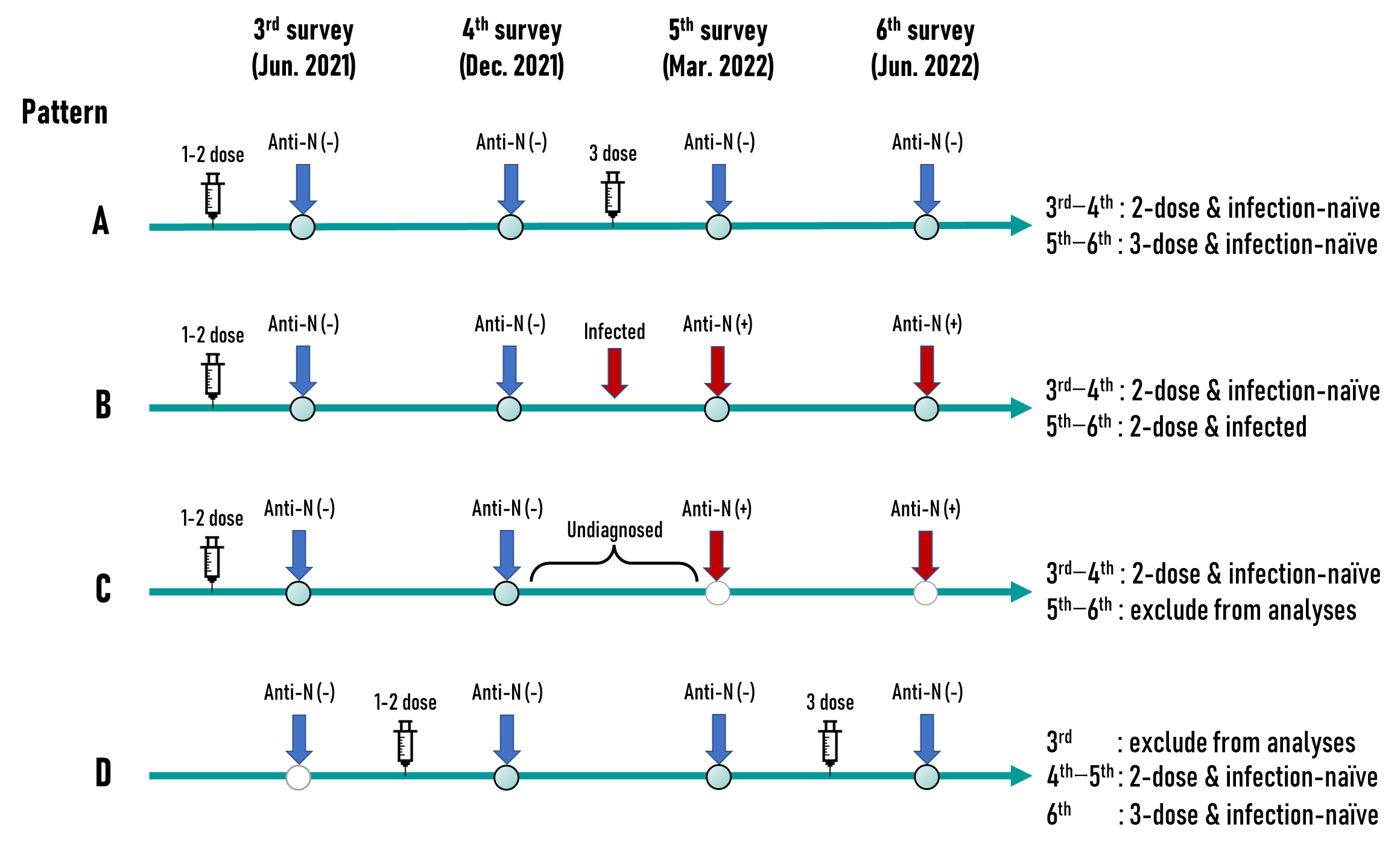

**Supplemental Figure 2.** Inclusion and exclusion patterns for analyses.

Anti-SARS-CoV-2 nucleocapsid protein (anti-N) positivity was tested with Abbott and Roche assays and was defined when either or both assays were positive.

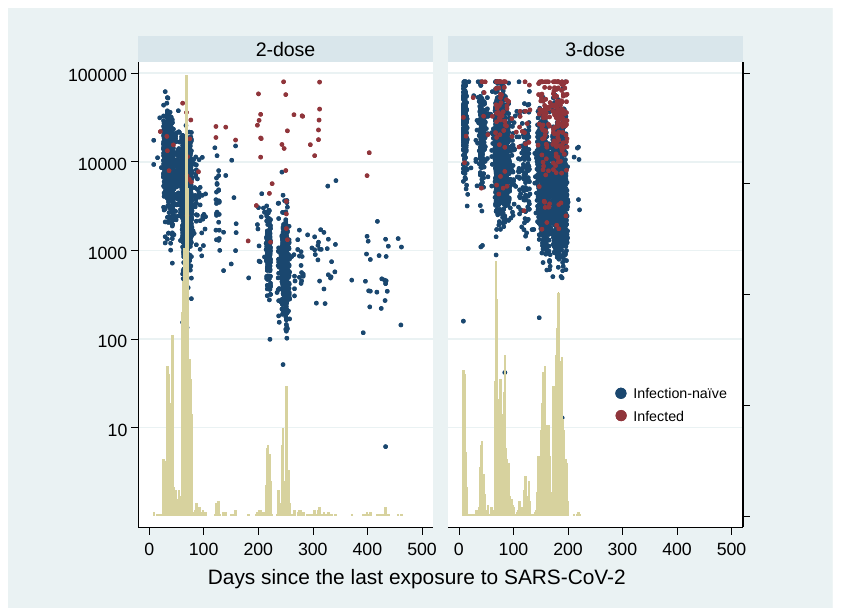

**Supplemental Figure 3.** Histograms of data at each time point and corresponding antibody titers after two (left) and three (right) vaccinations.

The median intervals (min–max) from the last SRAS-CoV-2 exposure (infection or vaccination) to the blood sampling among 2- and 3-dose recipients were 68 days (8–462) and 122 days (8–221), respectively.

**Table S1.** Comparison of the waning slope of antibody titers from the last immunological event across the sequence pattern of vaccination and infection.

| Sequence of vaccination and infection | **No.** | **Estimated spike antibody titers (AU/mL), GMT (95% CI)** | | | |  | **Slope of antibody waning, RoM (95%CI)** | | |
| --- | --- | --- | --- | --- | --- | --- | --- | --- | --- |
|  |  | **Day 14** | **Day 30** | **Day 90** | **Day 180** |  | **Per 30 days** | **Comparison of slopes** | |
| Infection-naïve | 6576 | 19335  (18570–20131) | 15040  (14574–15522) | 6467  (6292–6647) | 2436  (2356–2519) |  | 0.69  (0.69–0.70) | reference | – |
| Infection to vaccination | 52 | 28898  (18955–44057) | 22342  (15691–31812)* | 10339  (7620–14029)* | 5788  (4107–8157)* |  | 0.77  (0.71–0.82) | 1.10  (1.03–1.19) | reference |
| Vaccination to infection | 246 | 29253  (24740–34590)* | 29460  (26317–32979)* | 24700  (22065–27650)* | 10408  (7116–15224)* |  | 0.80  (0.73–0.87) | 1.15  (1.06–1.25) | 1.04  (0.93–1.17) |

*: P<0.05 (reference is infection-naïve)

Infection-naïve group includes those who received 2- or 3-dose without a history of COVID-19 and tested negative with anti-SARS-CoV-2 N assays.

Infection to vaccination group includes those infected before two doses and those infected before three doses (i.e., infected when immune-naïve status).

Vaccination to infection group includes those infected after two doses and those infected after three doses (i.e., infected when already having vaccine-induced immunity).

Shown are the estimated GMT of anti-SARS-CoV-2 spike protein antibody titers on 14, 30, 90, and 180 days since the last immune-conferring event (vaccination or infection) and the estimated RoM of antibody waning slopes, with adjustment for vaccination status (2- or 3-dose), an interaction between vaccination status and time, age (<40 or ≥40 years), sex (male or female), body mass index (<27.5 or ≥27.5 kg/m^2^), coexisting diseases (yes or no), immunosuppressant use (yes or no), smoking status (current or non-smoker), and frequency of alcohol drinking (<1 or ≥1 time/week).

GMT, geometric mean titer; RoM, ratio of means

**Table S2.** Comparison of the waning slope of antibody titers from the last immunological event across infected waves.

| Sequence of vaccination and infection | **No.** | **Estimated spike antibody titers (AU/mL), GMT (95% CI)** | | | |  | **Slope of antibody waning, RoM (95%CI)** | | |
| --- | --- | --- | --- | --- | --- | --- | --- | --- | --- |
|  |  | **Day 14** | **Day 30** | **Day 90** | **Day 180** |  | **Per 30 days** | **Comparison of slopes** | |
| Infection-naïve | 6576 | 19284  (18519–20080) | 15013  (14546–15495) | 6468  (6293–6648) | 2437  (2357–2520) |  | 0.69  (0.69–0.70) | reference | – |
| Infected during pre-Omicron waves | 71 | 34785  (23271–51996) | 26993  (19533–37301) | 12328  (9816–15482) | 6245  (4868–8012) |  | 0.75  (0.70–0.80) | 1.08  (1.01–1.15) | reference |
| Infected during Omicron BA.1/BA.2 waves | 254 | 29415  (24896–34775) | 29109  (26010–32577) | 23460  (20971–26246) | 10067  (6906–14676) |  | 0.80  (0.73–0.87) | 1.15  (1.05–1.25) | 1.06  (0.96–1.18) |

GMT, geometric mean titer; RoM, ratio of means

Pre-Omicron waves: February 2020 to December 2021.

Omicron BA.1/BA.2 waves: January 2022 to June 2022.

Shown are the estimated GMT of anti-SARS-CoV-2 spike protein antibody titers on 14, 30, 90, and 180 days since the last immune-conferring event (vaccination or infection) and the estimated RoM of antibody waning slopes, with adjustment for vaccination status (2- or 3-dose), an interaction between vaccination status and time, age (<40 or ≥40 years), sex (male or female), body mass index (<27.5 or ≥27.5 kg/m^2^), coexisting diseases (yes or no), immunosuppressant use (yes or no), smoking status (current or non-smoker), and frequency of alcohol drinking (<1 or ≥1 time/week).
